# “I think it would be better for those of us who have the disease not to be ashamed”. Insights from people living with chronic hepatitis B virus infection and healthcare workers providing HBV care in Kilifi, Kenya

**DOI:** 10.1101/2024.11.28.24317957

**Authors:** Louise O Downs, Juliet Odhiambo, Mwanakombo Zaharani, Oscar Chirro, Benson Safari, Janet Seeley, Philippa C Matthews, Nadia Aliyan, Nancy Kagwanja

**Affiliations:** Nuffield Department of Medicine, University of Oxford, UK; KEMRI-Wellcome Trust Research Programme, Kilifi, Kenya; London School of Hygiene and Tropical Medicine, Keppel St, London, WC1E 7HT, UK; Africa Health Research Institute, KwaZulu-Natal, South Africa; The Francis Crick Institute, 1 Midland Road, London NW1 1AT, UK; Division of Infection and Immunity, University College London, London, UK; University College London Hospital, 235 Euston Road, London NW1 2BU, UK; Kilifi County Hospital, Kilifi, Kenya

**Keywords:** Hepatitis B virus, Patient voice, Lived experience, Barriers to care, Care cascade, Africa, Kenya, Peer support, Education, Decentralisation, Treatment, Stigma

## Abstract

Chronic hepatitis B infection (CHB) causes over 1 million deaths annually, with a large burden of morbidity and mortality in the WHO-African Region (WHO-AFRO) where <5% of people are diagnosed and 0.2% are on treatment. Studies have shown that understanding of hepatitis B virus (HBV) here is often poor, and people living with HBV (PLWHB) can experience stigma and discrimination. However there has been little documentation on the impact of an HBV diagnosis on the lives of PLWHB in the WHO-AFRO region or community involvement in improving care provision. We undertook two focus group discussions (FGDs) with PLWHB and two with healthcare workers (HCWs) providing HBV care at Kilifi County Referral Hospital (KCRH), Kenya to explore experiences of living with HBV and barriers to accessing care. FGDs were conducted primarily in Kiswahili, transcribed verbatim and translated into English. The data were analysed thematically using NVivo version 14.

PLWHB and HCWs at KCRH had a good understanding of HBV which was likely influenced by a concurrent research study on HBV, however they reported low awareness in the general community, and there is no local name for the infection. Many PLWHB were shocked at their initial diagnosis with mixed reactions from friends and family. Costs of transport and concerns about lost employment were the biggest barriers to care. Many people suggested decentralised clinics would reduce loss to follow up, however others would rather be treated far from home to preserve anonymity. Stigma was highlighted as a major issue, leading to feelings of isolation, rejected and discrimination. Community education, wider testing and advocacy by well-respected community members were mentioned as key methods to reduce HBV transmission. Decentralisation of clinics may improve access to care; however, this needs to be developed in careful consultation with PLWHB to ensure they are acceptable and accessible to all.

## Introduction

Approximately 256 million people worldwide are living with chronic hepatitis B infection (HBV) with the highest burden of infection being in the WHO-African Region (WHO-AFRO) (1). HBV has, however, been neglected in terms of healthcare provision, education, research and policy (2,3). An estimated 4.2% of people living with HBV (PLWHB) in WHO-AFRO are diagnosed, and knowledge around HBV both in communities and healthcare facilities is often limited (1,4). In Kenya, an estimated 3-5% of the population is living with HBV (5), but screening is largely restricted to those living with HIV, blood donors, dialysis patients and in some locations antenatal screening. There is minimal population screening and little vaccination outside of the childhood immunisation series. Elimination efforts in Kenya are gaining traction with the Ministry of Health working on new HBV management guidelines and a National Strategic Plan; the community voice along with challenges faced by healthcare workers (HCWs) in caring for PLWHB must be considered in these efforts.

Throughout the WHO-AFRO region, reports are emerging of the impact of an HBV diagnosis on those living with the infection, and a new HBV stigma survey has been piloted in selected countries by the World Hepatitis Alliance (6). It is widely agreed that representation of those with lived experience is urgently needed to improve representation and advocacy, and inform improved service provision, which in turn will drive progress towards global elimination targets (7). The positive role of peer support workers in HBV care is being increasingly recognised, to encourage testing, improve treatment adherence and reduce stigma (8). Along with this, healthcare workers (HCWs) providing care for PLWHB have a unique insight into issues including misinformation, stigma and barriers to long-term follow-up and medication. They also understand the current healthcare infrastructure for PLWHB, have insights into gaps and challenges, and can advise how access could be improved.

We set out to engage people living with HBV, and HCWs caring for PLWHB in Kilifi County Referral Hospital (KCRH), Kenya, to generate evidence to inform care delivery, by establishing a platform for scale up of qualitative data collection in this setting and elsewhere. We convened focus group discussions (FGDs), aiming to i) explore understanding of HBV infection on an individual and community level, ii) discuss how the lives of PLWHB have changed since their diagnosis and iii) investigate barriers to receiving care and consider how these could be reduced. The methods and results of this study are reported in compliance with the Standards for Reporting Qualitative Research (SRQR)(9).

## Materials and methods

### i) Study context

We conducted FGDs with both PLWHB and HCWs providing HBV services. This study was conducted at Kilifi County Referral Hospital (KCRH), a referral facility in Coastal Kenya, servicing an area of around 12G000 km and a population of 1.5Gmillion people. KCRH has 300 inpatient beds and sees 193G000 outpatients every year. Table 1 summarises the distribution of health facilities in Kilifi County (10). Poverty levels in Kilifi are above the national average for Kenya, with 53% of the population living in poverty in 2022 compared to the national average of 40% (11). At the time of this study, HBV care at KCRH was provided at the Comprehensive Care Clinic (CCC), which is a secondary care outpatient clinic primarily designed for people living with HIV. The CCC provides adherence counselling, medication and follow up for both adults and children living with HBV. At KCRH, there is no routine mechanism for offering further laboratory or imaging assessment to determine HBV treatment eligibility, therefore standard care is that all those testing HBsAg positive receive nucleoside analogue (NA) therapy using tenofovir/lamivudine (TDF/3TC) combination therapy. This approach is in line with conditional recommendations for ‘treat all’ which are incorporated in new WHO guidelines released in 2024 (12).

**Table 1:** The distribution of health facilities in Kilifi County, Kenya (10).

| Health Facility Type | Number in Kilifi County |
| --- | --- |
| Dispensary | 132 |
| Health Centre | 18 |
| Public Hospitals |  |
| County Referral Hospital | 1 |
| Sub-County Hospital | 8 |
| Faith Based Facilities | 25 |
| Private, for-profit health facilities | 186 |

### ii) Data collection

We conducted four FGDs, two with PLWHB (consisting of six men and seven women) on 18 June 2024, and two with HCWs (consisting of seven men and seven women) on 14 November 2024. PLWHB were invited to participate in FGDs during peer support educational sessions initiated by the CCC matron when coming to pick up medication, and HCWs were invited via a group messaging platform for CCC staff. Participants were purposively sampled to create groups with diverse ages and experience, and in the case of HCWs to ensure a spread of cadres involved in HBV service delivery.

FGDs were held in private rooms in the CCC to promote participant privacy. Participants were given a number from 1-6 or 1-7 within their group to enhance anonymity of the respondents. FGDs with PLWHB were conducted primarily in Kiswahili by MZ, OC, and BS and facilitated by JO, a member of the Community Liaison Group from KEMRI Wellcome Trust Research Programme experienced in qualitative data collection. FGDs with HCWs were conducted in both English and Kiswahili by MZ and JO, facilitated by LD. Topic guides were used to direct FGDs, informed by study objectives and literature about patient experiences of accessing and receiving HBV care. The topic guides covered the understanding of HBV, reactions to diagnosis, the impact on the lives of PLWHB, barriers to care and how these could be reduced (13). FGDs took around 1.5 hours each, were audio recorded using an encrypted dictation device from KWTRP, and those participating were given refreshments. PLWHB attending specifically for the FGDs were compensated lost earnings of around USD $5, as is standard for community engagement activities at KWTRP.

### iii) STRIKE-HBV partner study

This work was undertaken in collaboration with the STRIKE-HBV study, a research study funded by the Wellcome Trust, based at KEMRI-Wellcome Trust Research Programme (KWTRP). STRIKE-HBV offered free HBV testing to all those attending KCRH between March 2023 – May 2024, and free liver health assessment to anyone living with HBV as previously described (14). STRIKE-HBV enrolled 102 PLWHB, including those newly diagnosed and those already known to be living with HBV and engaged in care. Activities undertaken by STRIKE-HBV including the status of HBV care in KCRH are detailed in table 2 and educational material developed for STRIKE-HBV is available online (15). All PLWHB who took part in these FGDs were also enrolled in STRIKE-HBV.

**Table 2:** Detailing i) Care for people living with HBV (PLWHB) at Kilifi County Referral Hospital (KCRH) prior to the STRIKE-HBV study; ii) Interventions undertaken by the STRIKE-HBV study and iii) Changes in care for PLWHB after STRIKE-HBV completion. Information leaflets were developed in collaboration with the Hepatitis B Foundation (16). CME sessions mostly targeted doctors, nurses, pharmacists and students. OPD – outpatients department; CCC – comprehensive care clinic.

| Category | Prior to STRIKE-HBV | Interventions undertaken by STRIKE-HBV | Situation after STRIKE-HBV |
| --- | --- | --- | --- |
| **HBV education for staff** | None provided.<br>Patients managed in the CCC with no staff training. | Hospital education sessions as part of continuing medical education (CME) and to individual departments, particularly the CCC. | Staff have continued HBV education including reaching out with questions.<br>KCRH is developing hospital specific guidelines for HBV management. |
| **HBV community education** | None provided. | - Health talks in Kiswahili detailing HBV symptoms and transmission 3 x week in KCRH OPD.<br>- Information leaflets provided (15)<br>- Education for PLWHB combined with peer support sessions at the CCC. | Staff able to provide improved education to PLWHB and antenatal women regarding HBV testing. |
| **HBV testing** | No routine HBV testing either free of charge or paid. No advocacy for testing. | Provision of free HBV testing for non-pregnant adults from March 2023 – June 2024. | HBV testing promoted and offered to antenatal women (although currently at an individual cost) |
| **Peer Support** | No peer support for HBV | Nil specific implemented by STRIKE-HBV, but study staff facilitated peer support sessions initiated by the CCC by providing education. | CCC began peer support sessions for PLWHB during STRIKE-HBV and these are continuing following study completion. |

### iv) Data Analysis

Following FGDs, audio files were transcribed verbatim and translated into English and analysed in Nvivo version 14. Both inductive and deductive thematic analysis (17) was used to scrutinise the data to identify and analyse patterns and draw out themes using constant comparison technique (18). Transcripts from all FGDs were examined on a line-by-line basis and the coding framework was first developed by LD and refined on discussion with the research team. Codes were developed based on the original questions asked by the interviewers and areas of interest to the authors, along with new themes being identified ‘a priori’ based on existing literature and tailored to the local context (4,19).

### v) Ethics and governance

This study was approved by the Scientific Ethics Review Unit (SERU) (SERU 3416). Written consent was obtained from all FGD participants prior to participation in FGDs, including consent for audio recording.

## Results

Focus groups with PLWHB included six men and seven women with a median age of 37 years (range 24 – 61). Five participants had been newly diagnosed with HBV in the STRIKE-HBV study, eight already knew their diagnosis prior to enrolment in STRIKE-HBV. Two of those participants who were newly diagnosed had family members already known to be living with HBV.

HCW focus groups consisted of nine women and five men, with a median age of 31 years (range 18 – 61). The range of cadres included three clinical officers, three community health promotors (CHPs), two nurses and then one of each: counsellor, adherence supervisor, peer educator, nutritionist, records officer, and laboratory technician.

### i) Knowledge and understanding of HBV

Both PLWHB and HCWs knew that HBV is a virus causing liver disease and is transmitted sexually or by contact with infected blood. Sexual transmission was mentioned most by both groups, including men and women, and still considered the most common route of transmission, especially by PLWHB, although other routes were mentioned. As participating PLWHB were those already receiving treatment, they were aware of HBV medication, and some HCWs specifically mentioned the use of tenofovir/lamivudine combination therapy. In both groups however, there was still some confusion around what happens after taking treatment for 6 months - some PLWHB thought they would be cured; however others knew this was not the case:

> *“In addition, if you have liver disease, you can, if you use the medicine properly for six months and if you are tested, it may be that the virus is gone”, (participant 5, PLWHB, male, FGD 1).*

Within the HCW groups, although there was discussion around poor community HBV knowledge, they also noted a significant knowledge gap amongst HCWs prior to the STRIKE-HBV study. They felt STRIKE-HBV had given them confidence in HBV management, and that many healthcare providers in other facilities were still providing incorrect information:

> *“At first, I didn’t know how to give the treatment. I used to just guess. I didn’t know how long I should be giving this medication. I didn’t even have the knowledge of package information to give the patient but right now I can stand comfortably with confidence and tell the patient what he or she is going to take and for how long the treatment should be and what he or she should do.” (participant 7, HCW, male, FGD 2)*

When discussing a local language description of HBV, participants from both groups stated it was an unknown disease, or only described by the symptoms:

> *“But this hepatitis B virus is unknown, let’s say it has no name. People don’t know, they only know AIDS”, (participant 1, PLWHB, female, FGD 2)*.

> *“In Kilifi they just describe the complications, when you have jaundice they say “yellow”, when you have ascites they say “mwamimba” [pregnancy], only that.” (participant 1, HCW, female, FGD 2)*

Others reported HBV infection was called *‘cancer’*. Several people mentioned that they had heard of HBV as it was causing illness in their village, and perceived it to be a death sentence:

> “*I knew it [HBV] was killing people in the village. You don’t know if he is starting [to get unwell] but you will only know that something has reached. That is, they don’t get medicine, that is, there is no nursing, it is only when he is taken to the hospital, when he begins to be treated, then death has already arrived” (participant 6, PLWHB, male, FGD 1)*.

Several respondents in both HCW and PLWHB groups mentioned that PLWHB were sometimes thought to be bewitched, and they would go to traditional healers to be cleansed and take herbal medications instead of tenofovir, or to church to pray to be cured:

> *“…those [with HBV] who have complications, or have ascites, in local language or native language we call it “mwamimba” [pregnant] and so they say that they have been bewitched and somebody has thrown something in their stomach and now like you are like a man but you are “pregnant”. (participant 5, HCW, female, FGD 2)*.

### ii) Comparison with HIV

Both PLWHB and HCWs discussed the confusion between HBV and HIV, particularly given that care is received in the same clinic, and the medication is the same. Many PLWHB found it difficult to accept they did not have HIV, and friends and family sometimes accused them of lying. This was common to both men and women. Many PLWHB felt they needed to ‘prove’ they didn’t have HIV by going with family or partners to be tested:

> *“…one day my wife came and explained to me, “I heard some people say that you have AIDS.” I told her, “No, I don’t have AIDS and if you want to know, let’s go, let’s all go and be tested” (participant 5, PLWHB, male, FGD 2)*.

> *“… [many] people who don’t understand and then once they get the results and then they are being escorted to this building CCC, majority don’t get to understand clearly the relationship like we are telling them it’s not HIV and we are giving them ARV’s (anti-retroviral therapy).” (participant 1, HCW, female, FGD 2)*.

Several times it was mentioned that HIV was better funded than HBV, and there was not the same level of resources for the two infections.

### iii) How lives have changed since HBV diagnosis

We initially discussed reactions to diagnosis, both individually and those of friends and family. Many PLWHB were shocked at their diagnosis and felt anxious as they had little knowledge of HBV. HCWs reported many newly diagnosed people thought they were dying, and associated HBV only with end stage disease. Counselling helped alleviate anxiety and was felt to be very important by both PLWHB and HCWs. Peer support sessions were motivating and HCWs thought these should be given more routinely to allow PLWHB to discuss their concerns. Several PLWHB mentioned that once they got used to their diagnosis, they realised they could continue to take the medication and live just like anyone else.

Many women diagnosed with HBV had been tested as a condition for travelling abroad for work, and their diagnosis had not been properly explained to them which caused a lot of distress. They felt lost and abandoned, unable to pursue their plans, whilst peers and colleagues went off on their travels.

> *“He [employment agent] refused to tell me because he said [the problem was] either liver or kidney. I told him, “Now there is no disease that is good and there is no disease which is bad, you can just explain it to me.” He said, “Just go to the doctor and get tested or go and get an x-ray and he [the other doctor] will explain more, but I won’t explain it to you.”” (participant 3, PLWHB, female, FGD 2)*.

Reactions of friends and family varied. Some were very supportive, went to get tested themselves and encouraged participants to continue medication, but some partners left after the diagnosis. Sometimes after an initial lack of understanding, relatives came to the CCC to seek more information about HBV and its consequences.

Almost all participants commented that their lives had changed since HBV diagnosis, sometimes in beneficial ways, some less so. One male participant had considerably cut his alcohol consumption due to concerns about his liver:

> “*Even that time I came to be tested, I came already drunk…I have been told “The liver’s job is this and this and this and when you eat, the food is processed like this and this and this in the stomach”… I saw that there is a very important knowledge…. I stopped drinking alcohol; I continue to take medicine. So I see there is a big change. That is, since I have been drunk all the time and this time, I can be sober, I’m grateful”, (participant 1, PLWHB, male, FGD 1)*.

Many PLWHB had experienced non-specific symptoms that had improved with treatment such as tiredness, stomach pains and swelling, nausea, itching, heartburn and constipation. Symptom improvement with treatment encouraged them to continue:

> *“I was given about five days [of medication]. I came back, I came and was given for two weeks, I came back. But since then, I see changes myself even in my body. I don’t get hurt a lot and I don’t get a lot of fatigue”. (participant 7, PLWHB, female, FGD 1)*.

> *“I was diagnosed with this disease, and I just saw that I was already dying. But when I got this treatment, I am fine now because even when I was sleeping and I could feel my stomach like water going this way, when I turned over, I could feel like water coming back this way and I said, “My stomach has started to swell too.” But this time when I started this treatment, that issue stopped and the anxiety of saying that I was dying because I had not received [education]. This time I have received [education]. Now I know that if I use these drugs properly, I can live normally.” (participant 5, PLWHB, male, FGD 1)*.

Many participants perceived that they had less freedom as they felt they needed to ensure they were home from work at a particular time and sometimes decided not to engage in social activities due to concerns about taking their medication:

> *“But since I use these drugs, I always tell my friends that I can’t walk at night. Because when it reaches eight o’clock, I have to take medicine” (participant 7, PLWHB, female, FGD 1)*.

### iv) Current provision of HBV care in Kilifi

We discussed with HCWs what care was available for HBV both in KCRH and further afield, and how this had changed since the STRIKE-HBV study. Adherence counselling and medication had always been available at KCRH free of charge for HBV, however as described above, HCWs reported inadequate knowledge about medication and a lack of confidence when prescribing. In Kilifi County, diagnosis was most often made in private facilities prior to travel abroad for work, and treatment was provided primarily at KCRH. Public peripheral facilities and private hospitals often referred patients to KCRH as it was known to have an established hepatitis clinic. Occasionally people would be able to pick up their medication from subcounty hospitals and smaller health centres, but only after education and service establishment by HCWs from KCRH:

> *“So, there were no services [no hepatitis clinic] there, but we had to recreate the services for her to get the drugs. So, some have opted to go to a smaller health centre, and some to two sub- county hospitals, and so we have some sites which are offering [HBV treatment] but the diagnosis was done here.” (Participant 7, HCWs, male, Group 2)*.

The HCWs however still perceived that peripheral facilities require more support to increase their confidence in providing HBV care. Many health centres and dispensaries had HBV medication available along with adherence counsellors as they cared for those living with HIV, yet patients who desired to get their treatment from their nearby facilities were sometimes still referred upwards to KCRH:

> *“…as much as they wish to go there, some facilities have limited knowledge on client follow up and now it becomes a challenge, but we have clients who would prefer to take medication from a nearby facility. I have had two who have been returned [to KCRH from peripheral facilities] for further management because the dispensary is not well equipped to manage hepatitis clients.” (FGD 2, HCWs, female, participant 1)*.

Since STRIKE-HBV, HCWs reported they were able to provide better informed psychosocial support for PLWHB. They also encouraged PLWHB to talk to partners and family about testing to try to reduce stigma. They reported encouraging PLWHB to be ambassadors and educate the community about HBV and some HCWs were sharing their improved HBV knowledge with colleagues in peripheral facilities to support them in providing HBV care.

### v) Barriers to HBV care and how these can be reduced

#### a. Economic Factors

Economic factors seemed to be the biggest barrier to receiving care. Firstly, the cost of transport to attend a distant hospital-based clinic was mentioned by both HCWs and PLWHB. To counter this cost, many people felt HBV medication should be available at the local dispensary:

> *“Because the rest of us are far away [from the hospital], but if we find our local dispensaries nearby, these local medicines, the way we are known, they are placed there. It will be easy even for us because we won’t have to pay fare”, (participant 2, PLWHB, female, FGD 1)*.

> *“…they come from far and first they have the transport issues and secondly they are living in abject poverty and so most of them they cannot make it to Kilifi and so at least if we have the resources [medication] and we dispense the resources [medication] nearer to them then it will be better.” (participant 7, HCWs, male, FGD 2)*.

Several PLWHB mentioned the need to plan to make sure they had fare to attend clinic. Along with fare, the timing of clinic appointments had little flexibility, and often the patients experienced long waiting times. This was difficult with employment, with men in particular mentioning they struggled to get days off work leading to lost earnings:

> *“So the challenge always arises because maybe your boss refuses to agree with you, maybe it [the clinic return date] can be balanced until you get the chance to come take medicine or a meeting like this. You may find that your boss telling you, “Now you might not get paid today, or we will remove this day from your rest days, we will cut it there”. (Participant 5, PLWHB, male, FGD 2)*.

The wish for decentralised clinics however was not universal. Several people in both the patient and HCW groups mentioned that (despite the potential higher cost) some PLWHB would prefer to go to a distant clinic (where they are not known) to reduce stigma, and sometimes asking for money for transport caused problems with questions about where they were going:

> *“…many of them still have stigma so they are thinking like if I am seen there, I will be isolated, so many prefer to go to those far away hospitals that no one knows, very few said they were referred to go to a nearby hospital.” (Participant 4, HCW, female, FGD 1)*.

> *“[If I ask a friend]: Help me with 200 shillings ($1.5) so that I can go somewhere and then I will give you back.” He asks, “Where do you want to go?” What do you want to do with it?” Now that’s where I see the difficulty.” (Participant 5, PLWHB, male, FGD 1)*.

#### b. Stigma and lack of education

Stigma was raised as an issue by all focus groups, both men and women, with participants highlighting instances of self-stigma and community stigma. Concerning self-stigma, PLWHB felt they were being talked about, and others were distancing themselves for fear of infection:

> *“And if the food itself I can’t eat with them at the same table, it is as if I will infect them, I should have my own bowl and sit aside”, (participant 3, PLWHB, female, FGD 1)*.

As noted earlier, HIV and HBV services are currently integrated in KCRH and all clients are seen together in the same clinic. PLWHB revealed tensions around this, with some participants feeling they should have a separate clinic from those living with HIV, partly due to stigma when seen entering the HIV clinic, but also due to long wait times when being seen together. Some people felt those living with HIV were prioritised, although there was also agreement that separating those with HBV and HIV could increase stigma, and they should all be together because they had more in common than differences:

> *“I see they should just be together because we cannot separate them [HIV patients]. We all say we are birds of the same colour, we are going in the same direction. It means that if we separate them, it is like we are bringing that stigma and we don’t want it to be like that”, (participant 2, PLWHB, male, FGD 2)*.

Education was seen by both PLWHB and HCWs as the best way to combat stigma, including continuing education for PLWHB through expert led peer support sessions, along with informing the immediate family and community. Several people mentioned initiatives on television or the radio to spread information about HBV, and respected community members such as pastors and chiefs to encourage testing and treatment. Several people in the HCW group felt the community health promotors (CHPs) could play a pivotal role in HBV sensitisation and treatment adherence. They mentioned how CHPs are funded to visit people living with HIV at home but not those living with HBV:

> *“I feel the CHP should also be brought on board on issues of Hepatitis B sensitisation and treatment adherence. Because you will get CHP bringing a referral or a patient who had defaulted treatment for [HIV treatment] but they will never do follow up for a client with Hepatitis.” (Participant 1, HCW, female, FGD 2)*.

#### vi) Reducing transmission

When asking PLWHB how to reduce HBV transmission, practical ways were discussed to reduce contact with the virus including safe sexual practices, and one participant mentioned wearing gloves before helping anyone who was injured. Reducing the sharing of instruments that spread HBV was felt to be important, particularly needles, razors and toothbrushes, and a few participants mentioned vaccination, but did not distinguish groups they felt could benefit from vaccination, or how vaccination should be offered.

Both PLWHB and HCWs felt that community awareness is paramount in stopping the spread of HBV, including reducing fears of testing by educating people that HBV can be managed well with medication, which is available free of charge, and is not a death sentence. Participants suggested testing could be promoted through TV and radio campaigns, by religious leaders and the Ministry of Health:

> *“So you find when people already know that [HBV can be dangerous], if the pastors also intervene, after explaining, now you will get a large group that say, “Uh, let me look at myself.” And there I think we will find many people will volunteer [for HBV testing]. You can’t go to someone’s house and grab his shirt and bring him here, no, it’s up to him to decide and say, “At my own free will, let me check.””, (participant 6, PLWHB, male, FGD 2)*.

Access to testing was also mentioned by several PLWHB, ensuring the community knows this is available, where to be tested and ideally testing being offered for free. Many HCWs and PLWHB discussed that HBV testing should be part of the routine screening that everyone has when they go to a hospital, like HIV. There was also discussion of the benefits of antenatal testing for HBV, and how this should be routine:

> *“… There should be free testing, random or all over, everyone to be reached. I can say here in Kilifi, the people of Kilifi are lucky because even I am lucky because of your research and the services that are provided here.” (participant 3, PLWHB, female, FDG 1)*.

> *“…perhaps if the pregnant mother it is her first time, she must come with her husband so that they can be tested for AIDS, and the likes. Now I was thinking it would also be good if this hepatitis B was placed in that category, when they come to be tested, if she has come to the clinic, being tested for the HIV virus, they should also test the Hepatitis B virus…”, (participant 6, PLWHB, male, FGD 2)*

Reducing stigma around an HBV diagnosis was felt to be key in reducing transmission. If those living with HBV felt able to reveal and discuss their diagnosis, they could also be the ones to spread awareness and educate their communities:

> *“I think it would be better for those of us who have the disease to not be ashamed. I explain that I was diagnosed with that disease [HBV] and that I use those drugs, I tell the people there to go to the hospital, not to wait until the organisation reaches the grassroots, it will be too late”, (participant 4, PLWHB, male, FGD 2)*.

A common theme that ran through all the discussion questions was lack of education. Improved HBV awareness and knowledge was seen as crucial in all areas including reducing stigma, reducing transmission, improving testing and diagnoses and improving access to care. Both PLWHB and HCWs highlighted this throughout as paramount to improve the lives of PLWHB in Kenya.

## Discussion

We set out to explore the experiences of both PLWHB and HCWs providing HBV care in Kilifi Kenya aiming to understand HBV knowledge, impact of HBV on people’s lives and barriers to accessing care. Our findings, summarised in Table 3, add to the limited literature exploring the lives of PLWHB in WHO-AFRO countries. Several studies have evaluated different mechanisms to try and improve HBV care, however very few have included the patient voice, or discussed with HCWs involved with patient care when planning interventions, making this a valuable addition to HBV literature (20,21).

**Table 3:**
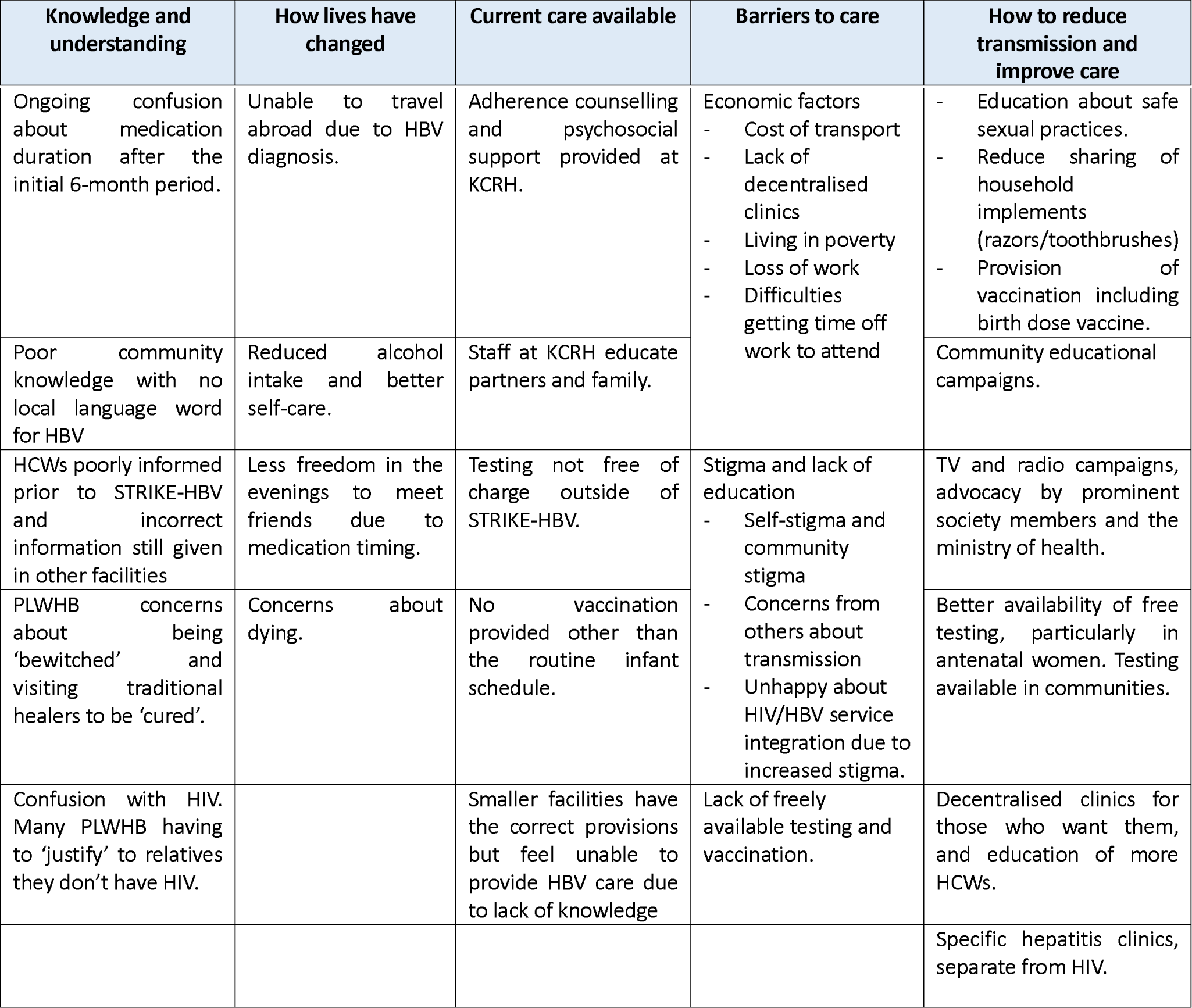
Summary of the different themes arising from focus group discussions (FGDs) with people living with hepatitis B virus infection (PLWHB) and healthcare workers (HCWs) caring for PLWHB at Kilifi County Referral Hospital (KCRH) in Kilifi, Kenya. HBV – hepatitis B virus.

Financial barriers have also been highlighted elsewhere (22–24), and can lead to catastrophic costs for individuals and families living with HBV (25). In our study, travel costs to the hospital clinic were seen as most prohibitive, whereas in studies from Burkina Faso and Ghana, costs of tests, monitoring and medications were mentioned more commonly (23,26). This may be because in our study testing and nucleoside analogue therapy were provided free at the point of care in KCRH which is not the case in many other countries. Many studies both from within WHO-AFRO and elsewhere have raised concerns around lost employment (27–29) as in our study, and this was particularly common amongst men. Lack of childcare was mentioned as a significant barrier to care in some studies outside of WHO-AFRO (28). This was not mentioned at all in our study or others from Ghana or Burkina Faso (23,24) possibly due to the different family structures in countries in WHO-AFRO compared to those in the Global North, with less reliance on parents being the sole care givers.

The concerns raised by our participants around the integration of HBV and HIV care particularly related to increased stigma has been discussed previously (30). The utilisation of existing HIV services is one of the key components hypothesised to enable decentralisation of HBV care. However, we suggest that caution is needed, and extensive community education and sensitisation should be done before this is implemented. We also found that there is often confusion between HIV and HBV, which was also the case in Burkina Faso where HBV was closely associated with HIV when explained to patients (23). Although this was seen as a negative association, the good understanding of HIV transmission routes did help explain HBV transmission. In Burkina Faso, those diagnosed with HBV outside of a hospital setting were often given poor information around HBV, similar to several of our PLWHB who were diagnosed pre-travel.

An interesting finding in our study was that many PLWHB felt they were restricted in the evenings due to taking medication, having to be home from work at a particular time or being unable to meet friends. This has been described in HIV literature as people are more likely to take their medication at home and do not want to carry tablets with them due to fear of discovery (31). It may also be that the HBV clinic has given instructions around when to take medication related to food to reduce side effects, and this is most convenient in the evenings. Education from the HBV clinic around the flexibility of medication timing could be helpful, so PLWHB understand it can take be taken at any time of day as long as this is consistent and does not need to interfere with any other social, domestic or work commitments.

Several suggestions from participants here on how to improve care are already included in national Kenyan guidelines on the management of HBV such as antenatal HBV testing and provision of vaccination to close family contacts (32), however these are not universally incorporated into care. To continue strengthening HBV care at KCRH and across the County, ensuring education is a funding priority is crucial. Education was seen as one of the most important initiatives to increase diagnoses, drive down stigma and improve HBV care pathways. As HCWs frequently transfer between facilities, even localised educational initiatives have the potential to disseminate knowledge more broadly.

## Caveats and limitations

We only included PLWHB in this study who were already linked to care and receiving treatment, and both HCWs and PLWHB had been involved in previous research and have engaged with a peer support group. These groups are likely to have better knowledge and PLWHB be more engaged in care than those not involved in such a study. Here we included people from a limited geographical area and only a small number of HCWs and PLHWB. To fully understand care barriers, we need to undertake similar discussions with PLWHB who have been lost to follow up, those not offered treatment and include community groups not tested or living with HBV; ideally a future study would expand to include PLWHB and HCWs accessing healthcare services both in Kilifi and further afield.

## Conclusions

Community education and decentralisation of care for PLWHB should be prioritised to reduce stigma, enable easier access to care, reduce travel times and costs to reduce health inequities and support wider roll-out of treatment. Community co-design is critical to ensure interventions are appropriate, accessible, acceptable and affordable, and can be sustained over time.

## Data Availability

Full recordings and transcripts in both English and Kiswahili of these FGDs are held on a secure server at KWTRP but could be accessed by other studies on request to and approval by the data governance committee.

http://dx.doi.org/10.6084/m9.figshare.28350341.v1

## Author Contributions

Study conceptualisation and design: LD, JO, MZ, OC, BS, PM

Data curation: LD, JP, MZ, OC, BS, NA, NK

Data analysis: LO, PM, JS, NK

Supervision: JS, PM, NA, NK

Writing – original draft and preparation: LD

Writing – Review and editing: JS, JO, PM, NA, NK

## Acknowledgements

We would like to thank all the PLWHB and HCWs who took part in these FGDs, along with the CCC team for facilitating the use of their clinic space to undertake this work. Without their enthusiasm and dedication to improving care for those living with HBV, this would not have been possible.

## Data Availability

Full recordings and transcripts in both English and Kiswahili of these FGDs are held on a secure server at KWTRP but could be accessed by other studies on request to and approval by the data governance committee. For the purpose of Open Access, the author has applied a CC-BY public copyright license to any author accepted manuscript version arising from this submission. This manuscript was written with the permission of Director KEMRI CGMRC.

